# Continuous vital sign monitoring of individuals with acute Lassa fever using wearable biosensor devices

**DOI:** 10.1101/2024.08.29.24312749

**Authors:** Brady Page, Raphaëlle Klitting, Matthias G. Pauthner, Steven Steinhubl, Stephan Wegerich, Margaret Kaiser, Foday Alhasan, Edwin Konuwa, Veronica Koroma, Ibrahim Sumah, Jenneh Brima, Tiangay Kallon, Brima Jusu, Sia Mator-Mabay, Isata Massaquoi, Mohamed Kamara, Fatima Kamara, Emilia Jaward, Angella Massally, Zainab Kanneh, Michelle McGraw, John Schieffelin, Donald Grant, Kristian G. Andersen

## Abstract

**Background:** Lassa fever is a fulminant viral illness associated with high in-hospital mortality. This disease constitutes a serious public health concern in West Africa, in particular Nigeria and the Mano River Union region (Guinea, Liberia, and Sierra Leone). In Sierra Leone, continuous monitoring of critically ill patients is hindered by a lack of equipment and personnel.

**Methods:** We used wearable biosensor devices to remotely monitor hospitalized individuals with acute Lassa fever in order to describe vital sign trends that may be associated with clinical outcome and to evaluate the feasibility of this approach in a resource-limited setting.

**Results:** The case fatality rate among participants (n=8) was 62.5%, with a median time from hospital admission to death of 2 days. Our results show that individuals who died (n=5) had higher mean heart rate (126 beats per minute) and respiratory rate (29 breaths per minute), as well as lower mean heart rate variability (10 ms), compared to those that survived (63 beats per minute, 22 breaths per minute, and 59 ms, respectively). Non-survivors also spent a greater proportion of their monitoring period in the age-specific tachycardia range (45.8%) compared to survivors (1.7%).

**Conclusions:** Although real-time monitoring of vital signs using wearable biosensors may have the potential to identify decompensations earlier than traditional bedside vital sign collection in a resource-limited setting, technical improvements are still needed to enable widespread use of this tool, for both clinical and research purposes.

## INTRODUCTION

The capacity of low- and middle-income countries to monitor and care for critically ill patients infected with high-consequence pathogens remains far below that of high-income countries where personnel, equipment, and specialized expertise are more accessible^1^. Previous efforts have addressed this disparity by employing innovative new technologies with the potential to economically and reliably begin to bridge this gap^2,3^.

Mobile health technologies–such as wearable biosensor devices for continuous vital sign monitoring–have the advantage of minimizing exposure to infectious patients while providing an abundance of objective data without much overhead or expertise, compared to more durable telemetry systems^4,5^. The promise of wearable sensors is that the early detection of certain physiological trends can warn clinicians about patients at risk for deterioration and lead to earlier intervention^6^. These devices have repeatedly been validated for remote vital signs monitoring and mortality prediction among unstable patients in low-resource settings, including during the 2014-2016 outbreak of Ebola Virus Disease in Sierra Leone^2,7–9^.

Lassa fever is another fulminant systemic viral illness which is endemic to Sierra Leone and is associated with annual outbreaks of high mortality^10^. A major driver of mortality in Lassa fever is multiorgan failure from hemodynamic collapse^11^. Sierra Leone, one of the world’s poorest and least developed countries, lacks the material and nursing resources to frequently and safely monitor patients with Lassa fever while maintaining rigorous containment measures ^12^.

In this study, we used wearable biosensor devices to continuously and remotely monitor individuals with Lassa fever in an under-resourced endemic area of Sierra Leone, in order to describe vital sign changes that accompany clinical outcomes and to evaluate the feasibility and efficacy of this approach in such a setting.

## METHODS

### Study design and setting

This prospective observational study took place in 2019-2021 at Kenema Government Hospital (KGH), the national treatment center for Lassa fever in Sierra Leone where all identified and suspected cases of the disease are transferred. KGH is a secondary referral center with a large catchment area that includes both urban and rural settings in the country’s Eastern Province.

### Patient selection and continuous physiological monitoring

Individuals of all ages admitted to KGH with acute Lassa fever between January 2019 and July 2021 were invited to enroll. Participants satisfied the case definition criteria for Lassa fever (Figure 1) and produced a positive serum antigen test for Lassa virus (Zalgen Labs, Frederick, MD, USA). Exclusion criteria included a history of allergy to skin adhesive. All confirmed cases were cared for in a designated Lassa ward with round-the-clock specialized nursing and enhanced isolation measures. Manual vital sign collection, diagnostic testing, and treatment were conducted throughout admission to the local standard of care. All patients admitted to the Lassa ward underwent rapid testing for malaria, received antibiotics as determined by the treating physician, and began treatment with intravenous ribavirin given as a 30 mg/kg loading dose, followed by 15 mg/kg every six hours for four days and 7.5 mg/kg every six hours for six more days.

**Figure 1.**
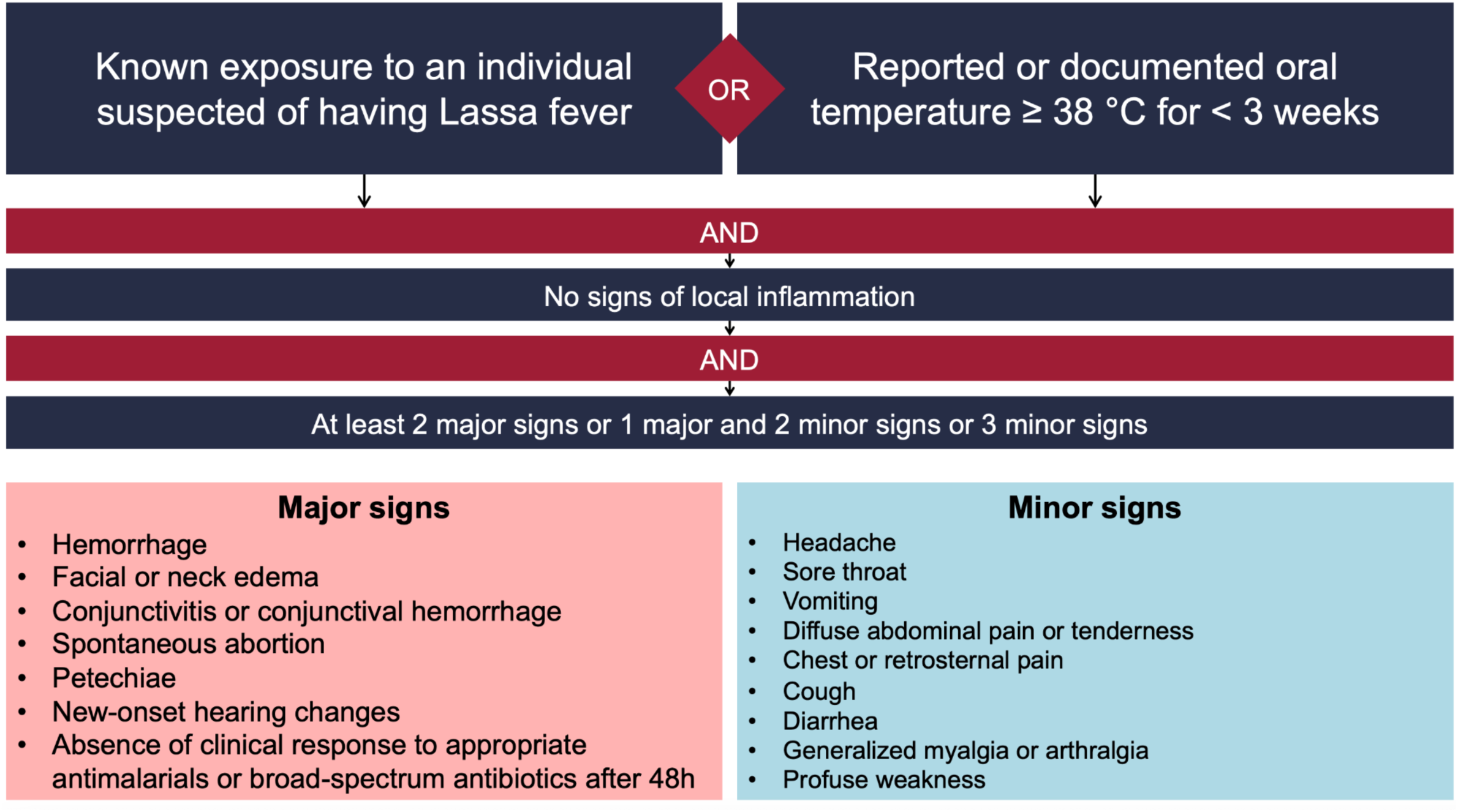
Lassa fever case definition criteria.

Upon admission to the Lassa ward, a battery-powered VitalPatch wearable biosensor device (VitalConnect, San Jose, CA, USA) was applied to each patient’s chest by a healthcare worker in full personal protective equipment (PPE). The device is an FDA-approved, 115 x 36 x 8 mm, 11 g, battery-powered, flexible strip with an adhesive side that attaches to the skin of a patient’s chest. Devices were linked via Bluetooth to Android devices running the physIQ data analytics platform (physIQ, Chicago, IL, USA), which were located in a nursing station outside of the isolation area. Given its observational nature, continuous vital signs collected as part of the study were not available to influence clinical care. All nurses received training in device placement, use of the analytic application, study procedures, and research ethics. Devices were worn until discharge from the Lassa ward, which was considered once patients had clinically improved, completed a 10 day course of ribavirin, and produced a negative serum antigen test for Lassa virus. If a device’s battery expired prior to discharge or there were issues with adhesion, a new device was applied.

### Physiological analysis

Waveform data from the EKG leads and thermistor present on biosensor devices was analyzed by the machine learning algorithms of the physIQ platform to generate the following parameters at one minute intervals: skin temperature, heart rate (HR), respiratory rate (RR), time domain heart rate variability (HRV), and EKG quality^7^.

### Statistical analysis

Descriptive statistics were used to summarize patient demographics and vital signs. Testing for statistical significance was not performed due to the study’s small sample size.

As a quality control measure, patients who did not produce at least twelve hours of continuous physiological monitoring data were not included in the analysis. Due to the presence of some spurious readings, patient physiological data was filtered to remove one minute data windows where EKG quality was less than 100%, skin temperature was < 33°C or > 42°C, or respiratory rate was 0. When comparing mean HR and RR values between survivors and non-survivors, only data up to 104 hours of monitoring were used, since up to this point there are multiple patients from both the survivor and non-survivor groups. For HRV, only data up to 83 hours of monitoring was included.

### Ethics statement

Written informed consent was obtained from all participants or, if an individual was unable to provide informed consent, from an appropriately identified surrogate. Written informed consent was obtained from the parent or guardian of each participant under 18 years of age. This study was approved by research ethics boards at Tulane University (IRB #140674) as well as the Sierra Leone Ethics and Scientific Review Committee.

## RESULTS

Thirty patients were admitted to the KGH Lassa ward during the study period. Twenty-two individuals (73.3%) were excluded for not yielding at least twelve hours of continuous physiological monitoring data, leaving eight participants to be included in the analysis. Discontinuous monitoring was due to both a lack of adhesion of the device, as well as bluetooth and cellular connectivity issues. Among the eight patients included, the median age was 6.5 years (0.4-40 years), 50% were female, the median time from onset of symptoms to admission was 7 days (2-21 days), and rapid malaria test was positive in 50% of participants. The in-hospital mortality rate was 62.5%, with a median time from hospital admission to death of 2 days. Patient demographics, presenting symptoms, vital signs on admission, and clinical outcome are presented in Table 1.

**Table 1.**
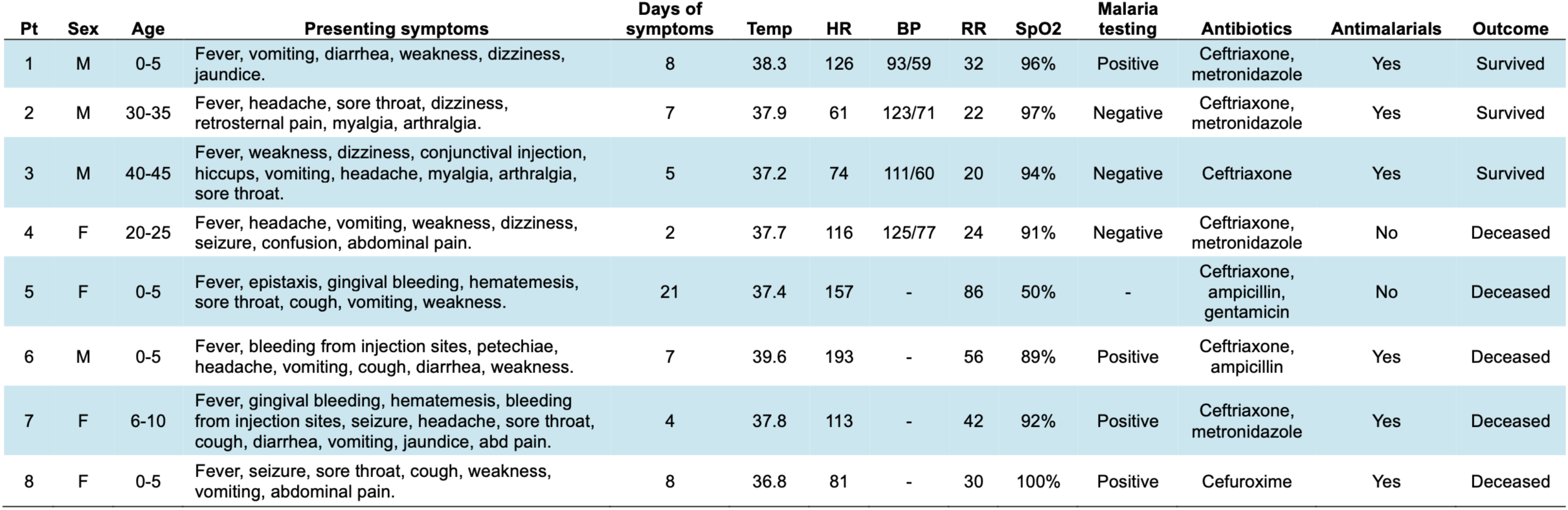
Baseline demographics and vital signs of participants on admission to the Lassa ward. Vital signs were collected manually on admission to the Lassa ward. Age is in years, temperature is in °C, heart rate (HR) is in beats per minute, blood pressure (BP) is in mmHg, respiratory rate (RR) is in breaths per minute, and peripheral oxygen saturation (SpO2) is expressed as a percentage. Days of symptoms are those prior to admission. Antibiotics are only those begun on admission.

| Pt | Sex | Age | Presenting symptoms | Days of symptoms | Temp | HR | BP | RR | SpO2 | Malaria testing | Antibiotics | Antimalarials | Outcome |
| --- | --- | --- | --- | --- | --- | --- | --- | --- | --- | --- | --- | --- | --- |
| 1 | M | 0-5 | Fever, vomiting, diarrhea, weakness, dizziness, jaundice. | 8 | 38.3 | 126 | 93/59 | 32 | 96% | Positive | Ceftriaxone, metronidazole | Yes | Survived |
| 2 | M | 30-35 | Fever, headache, sore throat, dizziness, retrosternal pain, myalgia, arthralgia. | 7 | 37.9 | 61 | 123/71 | 22 | 97% | Negative | Ceftriaxone, metronidazole | Yes | Survived |
| 3 | M | 40-45 | Fever, weakness, dizziness, conjunctival injection, hiccups, vomiting, headache, myalgia, arthralgia, sore throat. | 5 | 37.2 | 74 | 111/60 | 20 | 94% | Negative | Ceftriaxone | Yes | Survived |
| 4 | F | 20-25 | Fever, headache, vomiting, weakness, dizziness, seizure, confusion, abdominal pain. | 2 | 37.7 | 116 | 125/77 | 24 | 91% | Negative | Ceftriaxone, metronidazole | No | Deceased |
| 5 | F | 0-5 | Fever, epistaxis, gingival bleeding, hematemesis, sore throat, cough, vomiting, weakness. | 21 | 37.4 | 157 | - | 86 | 50% | - | Ceftriaxone, ampicillin, gentamicin | No | Deceased |
| 6 | M | 0-5 | Fever, bleeding from injection sites, petechiae, headache, vomiting, cough, diarrhea, weakness. | 7 | 39.6 | 193 | - | 56 | 89% | Positive | Ceftriaxone, ampicillin | Yes | Deceased |
| 7 | F | 6-10 | Fever, gingival bleeding, hematemesis, bleeding from injection sites, seizure, headache, sore throat, cough, diarrhea, vomiting, jaundice, abd pain. | 4 | 37.8 | 113 | - | 42 | 92% | Positive | Ceftriaxone, metronidazole | Yes | Deceased |
| 8 | F | 0-5 | Fever, seizure, sore throat, cough, weakness, vomiting, abdominal pain. | 8 | 36.8 | 81 | - | 30 | 100% | Positive | Cefuroxime | Yes | Deceased |

Among the eight participants included in the study, a total of 788.5 hours of continuous waveform data were collected, with 172.5 hours (21.9%) discarded for poor quality (Figure 2). Ultimately, 616 hours of waveform data were analyzed, with an average of 77 hours (12.7 - 151.7 hours) per patient (Figure 3). Four (50%) participants required the application of a new biosensor during their hospitalization: one when the battery on their first device expired and three more when there were issues with the device’s adhesive surface (Table 2). There were no adverse reactions to the devices.

**Figure 2.**
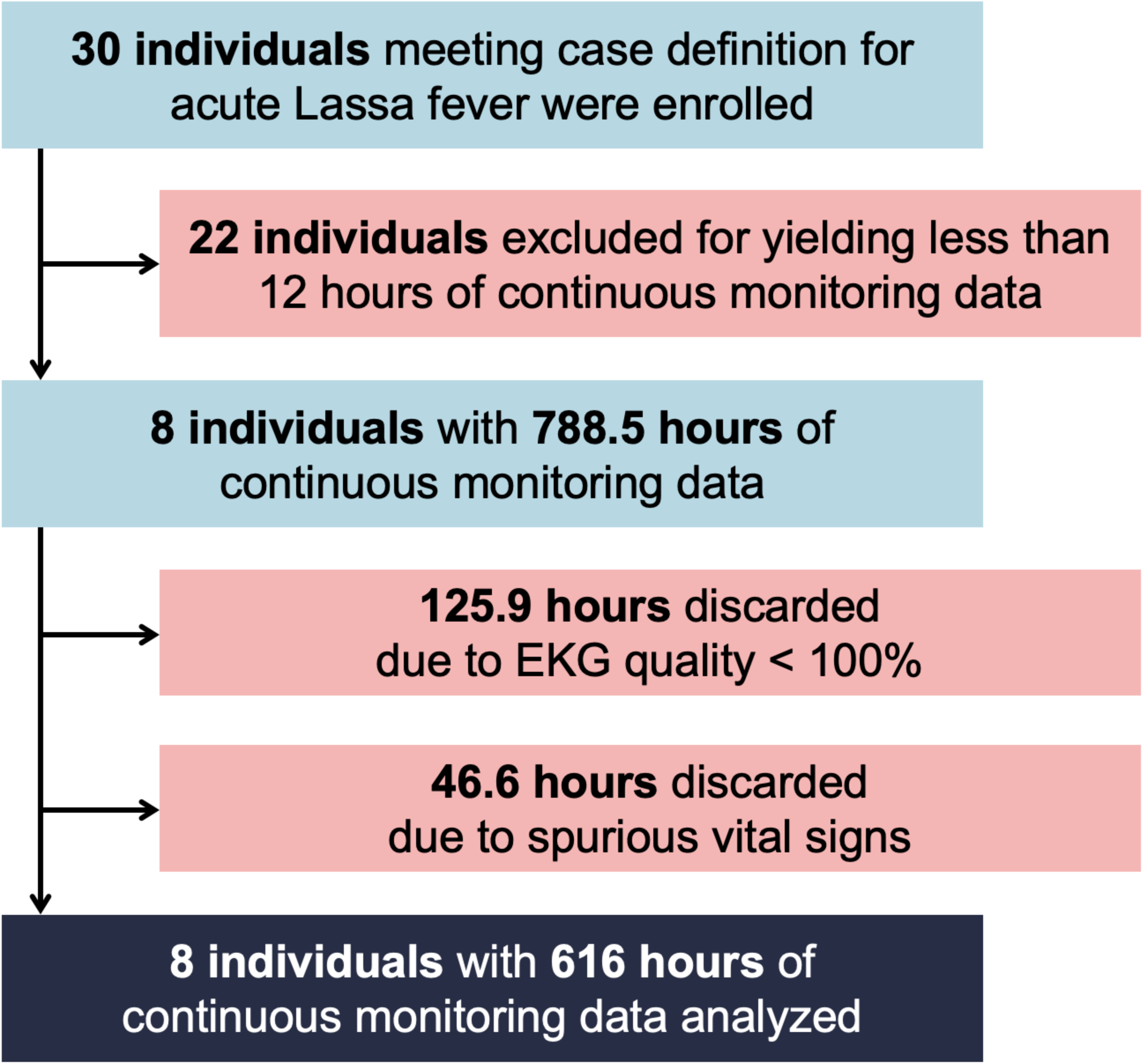
Flow chart of individuals and total hours excluded from analysis. A total of 30 patients were admitted to the KGH Lassa ward during the study period and enrolled in our study to be continuously monitored. Twenty-two individuals were excluded for yielding < 12 hours of continuous physiological monitoring data. Among the 8 participants included in the study, a total of 788.5 hours of continuous waveform data were collected, with 172.5 hours discarded for poor quality, defined as EKG quality < 100% or or spurious vital signs, which were defined as respiratory rate of 0 or skin temperature that was either < 33°C or > 42°C. Ultimately, 616 hours of waveform data were analyzed.

**Figure 3.**
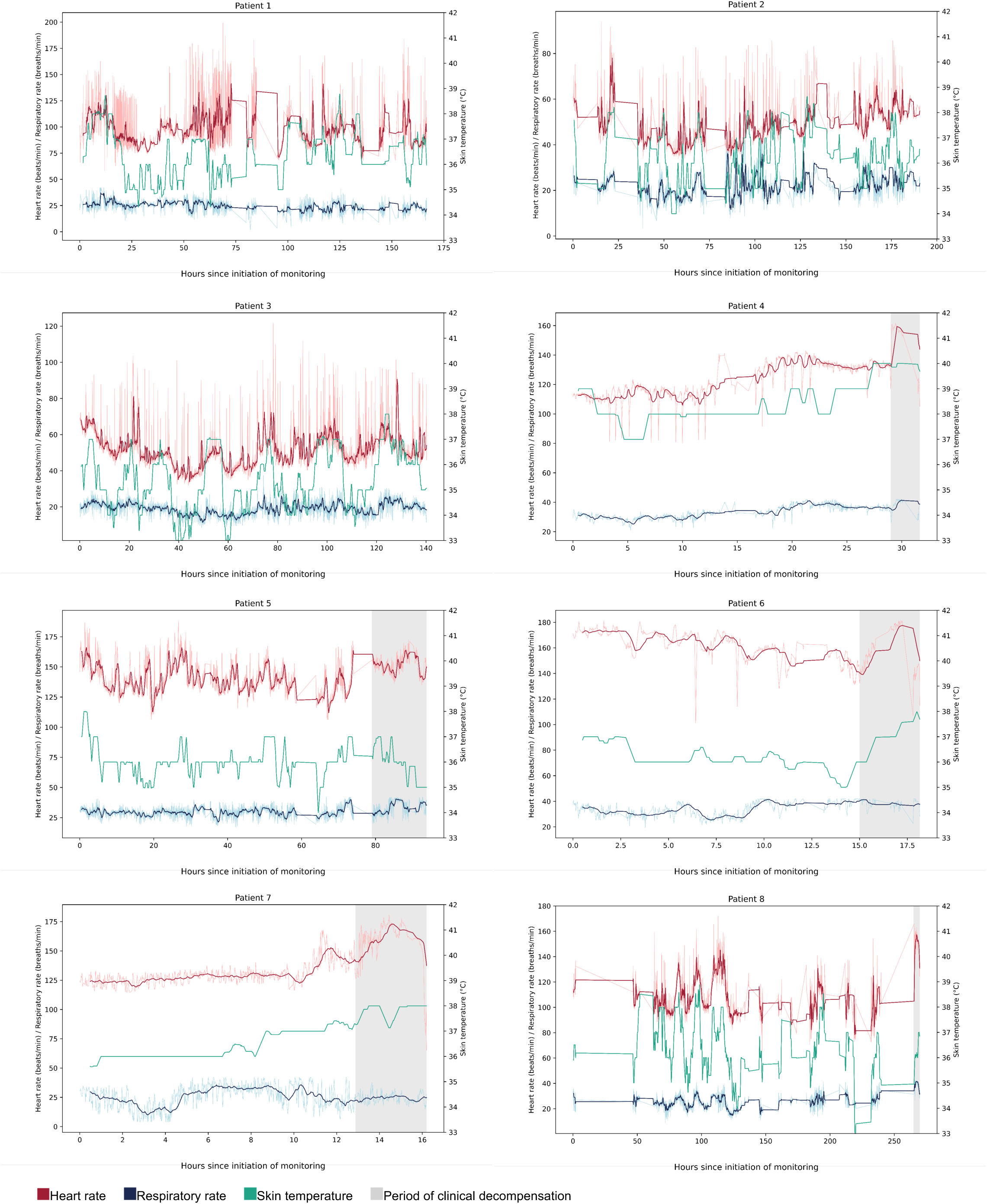
Heart rate, respiratory rate, and skin temperature during the course of acute Lassa fever. Trends are displayed for all eight individuals that were included in the analysis. The dark red and dark blue curves represent the thirty-minute rolling averages for HR and RR, respectively, while the light red and light blue curves represent HR and RR at one-minute intervals, as they were recorded by the device. Skin temperature is displayed as a thirty-minute rolling average.

**Table 2.**
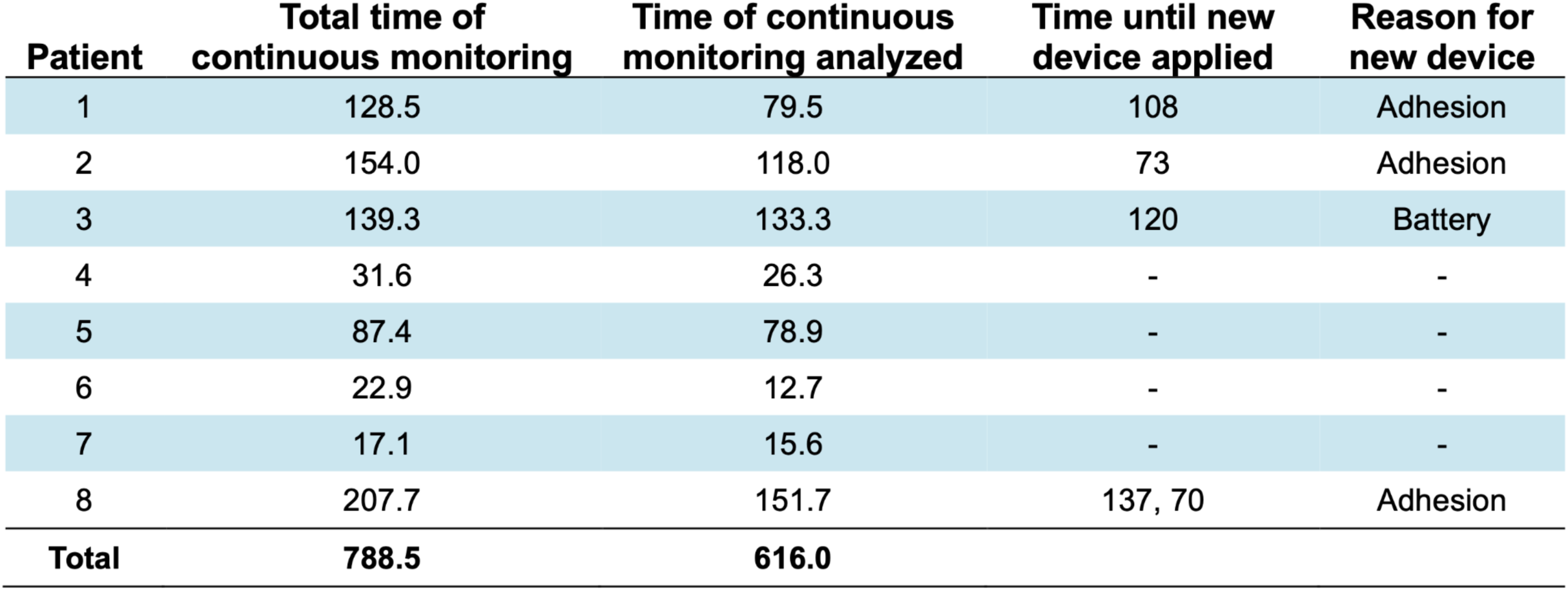
Features of continuous remote monitoring. Time is reported in hours.

| **Patient** | **Total time of continuous monitoring** | **Time of continuous monitoring analyzed** | **Time until new device applied** | **Reason for new device** |
| --- | --- | --- | --- | --- |
| 1 | 128.5 | 79.5 | 108 | Adhesion |
| 2 | 154.0 | 118.0 | 73 | Adhesion |
| 3 | 139.3 | 133.3 | 120 | Battery |
| 4 | 31.6 | 26.3 | - | - |
| 5 | 87.4 | 78.9 | - | - |
| 6 | 22.9 | 12.7 | - | - |
| 7 | 17.1 | 15.6 | - | - |
| 8 | 207.7 | 151.7 | 137, 70 | Adhesion |
| **Total** | **788.5** | **616.0** |  |  |

Among those that survived acute Lassa fever (n=3), mean HR and RR were lower than that of those who died (n=5) at nearly all time points over the first 104 hours of continuous monitoring (Figure 4). The mean HR and RR for survivors were 63 beats per minute and 22 breaths per minute, respectively, and 126 beats per minute and 29 breaths per minute for those that died. To account for age differences among our cohort–which was composed of both children and adults–we compared study HRs to established age-specific normal resting heart rates. For each of the three survivors, mean heart rates were below the age-specific thresholds for tachycardia; however, mean heart rates were in the tachycardic range for four (80%) of the non-survivors, whose heart rates were on average 9.4% higher than each’s age-specific threshold for tachycardia^13^. Overall, survivors spent 1.7% of their monitoring period in the age-specific tachycardic range, while non-survivors spent 45.8% of the monitoring period in the age-specific tachycardic range.

**Figure 4.**
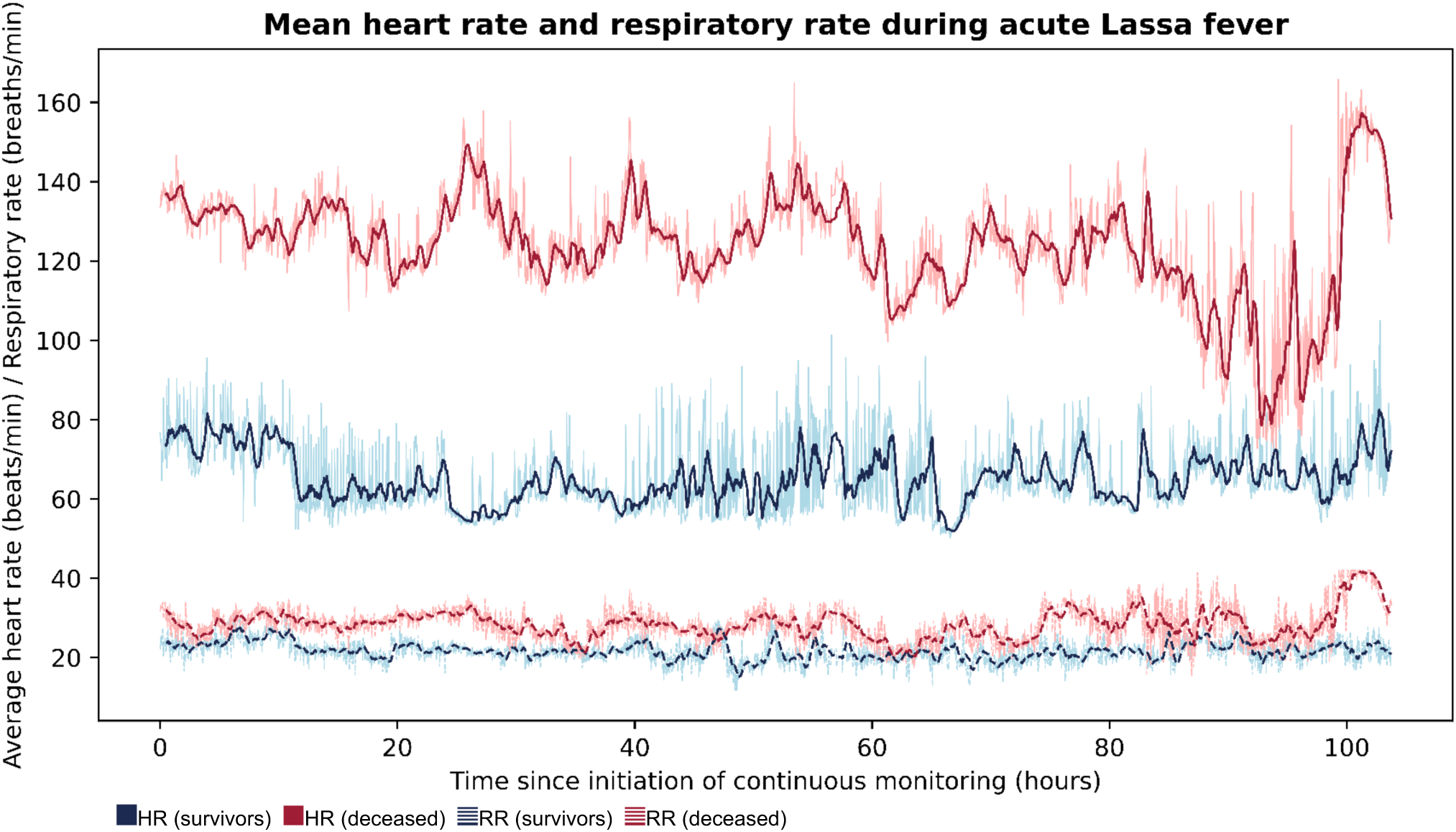
Mean heart rate and respiratory rate during acute Lassa fever among survivors and the deceased. The dark blue and dark red curves represent the thirty-minute rolling averages of vital signs for survivors (n=3) and the deceased (n=5), respectively, while the light blue and light red curves report vital signs at one-minute intervals. Solid lines represent HR and dashed lines represent RR. Among those that survived acute Lassa fever (n=3), mean heart rate (HR) and respiratory rate (RR) were lower than that of those who died (n=5) at nearly all time points over the first 104 hours of continuous monitoring. The mean HR and RR for survivors were 63 beats per minute and 22 breaths per minute, respectively, and 126 beats per minute and 29 breaths per minute for those that died.

Mean HRV among survivors was higher than that of those who died at nearly all time points over the first 83 hours of continuous monitoring (Figure 5). The mean HRV of survivors and non-survivors over the entire course of their monitoring periods were 59 and 10 milliseconds (ms), respectively. The mean skin temperature for survivors was 35.9 °C and for non-survivors was 36.5 °C.

**Figure 5.**
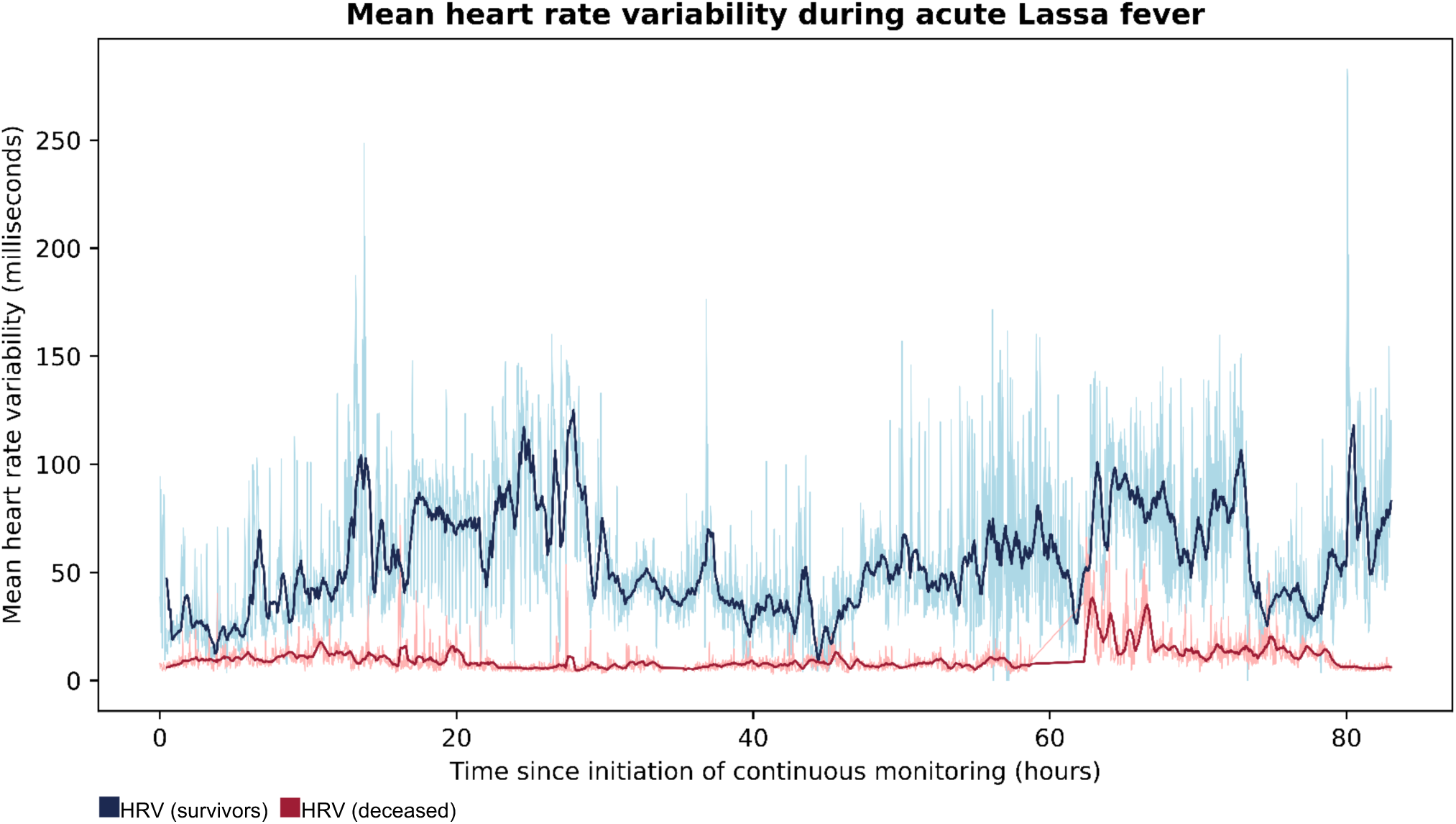
Mean heart rate variability during acute Lassa fever among survivors and the deceased. Time domain heart rate variability (HRV) as measured by the standard deviation from the mean time of RR intervals. The dark blue and dark red curves represent the thirty-minute rolling averages of vital signs for survivors (n=3) and the deceased (n=5), respectively, while the light blue and light red curves report vital signs at one-minute intervals. Among those that survived acute Lassa fever, mean HRV was higher than that of those who died at nearly all time points over the first 83 hours of continuous monitoring. The mean HRV of survivors and the deceased were 59 and 10 milliseconds, respectively.

Some interesting physiological phenomena were captured by the biosensors, both in real-time on the Android monitoring devices and retrospectively on the analytics platform. For example, over the course of their monitoring, Patient 4 is generally tachycardic with several episodes of HR returning to normal range (Figure 3). Visualization of the patient’s biosensor waveforms demonstrate the elevated HR with a high predicted probability of atrial fibrillation, which was due to frequent detection of ectopy (Figure 6). The patient then experiences multiple episodes of activity, which initially increase the HR before lowering it and reducing the probability of atrial fibrillation to zero. This interplay of heart rate, arrhythmia, and patient activity suggests coughing fits with subsequent increases in vagal tone that temporarily suppress atrial ectopy or ventricular responsiveness^14^.

**Figure 6.**
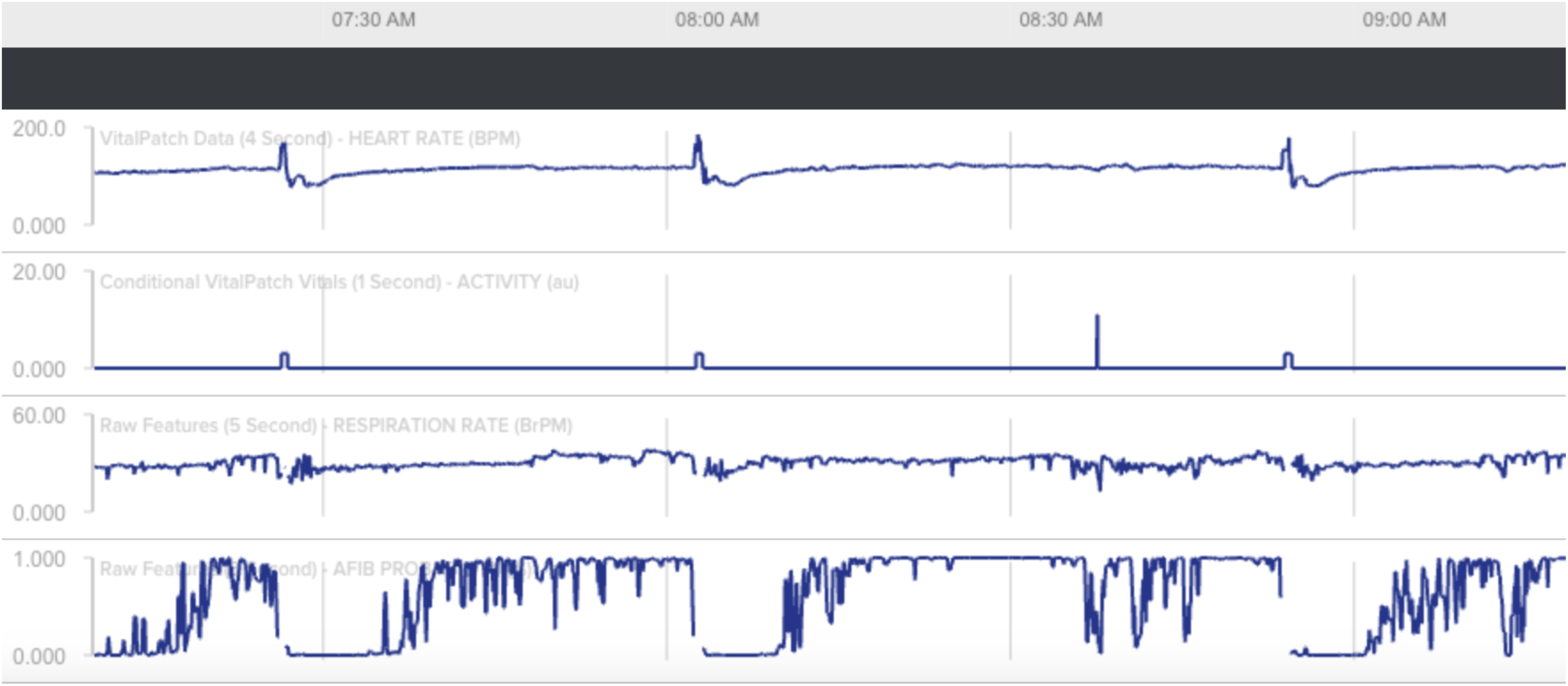
Analytic platform interpretation of waveforms produced from biosensor device data. An example of physiological parameters for Patient 4 changing in concert with one another. The patient has an elevated HR with a high probability for atrial fibrillation, as predicted by the analytics platform. The patient demonstrates several episodes of activity (positional changes from lying to sitting or standing), which initially increase the HR before lowering it and reducing the probability of atrial fibrillation to zero for periods of time. These interrelated findings may represent coughing fits with subsequent increases in vagal tone that suppressed atrial ectopy or ventricular responsiveness. Top waveform: heart rate (beats per minute). Second waveform: patient activity. Third waveform: respiratory rate (breaths per minute). Bottom waveform: probability of atrial fibrillation.

## DISCUSSION

This is the first study evaluating the use of wearable biosensor devices for the continuous physiological monitoring of individuals with acute Lassa fever. Although this study was not powered to make conclusions about physiological parameters and their associations with clinical outcomes, we did observe that patients who did not survive their illness had a higher mean HR and RR compared to those who survived. These findings are similar to what has been observed in similar settings for patients with bacterial sepsis^15^. It is worth noting that four of the five non-survivors were pediatric patients, while only one of the three survivors was a child, who are known to have higher resting heart rates than adults^16^. We attempted to account for this by using established age-specific thresholds to define tachycardia. Elevations in respiratory rate during Lassa fever could be explained by several potential etiologies including encephalopathy, anxiety, hypoxemia, pleural effusion, acute anemia, lactic acidosis from impaired tissue perfusion, or acidosis from renal failure^17,18^.

There is robust clinical evidence that alterations in HRV correlates with severity of systemic infection and has prognostic value as a predictor of multiorgan dysfunction and death^6,19–21^. A normal HRV is between 19-75 ms. On average, individuals who survived acute Lassa fever demonstrated HRV within the normal range (mean HRV 59 ms), while those who died were characterized by HRV below the normal range (mean HRV 10 ms)^22^. Increases in HRV over the course of infection have also been shown to correlate with survival in ICU patients with septic shock^23^. In our study, non-survivors failed to mount any sustained increases in HRV. Pre-admission measurements for all patients were unavailable, which precluded the opportunity for comparison of HRV during acute illness with individuals’ healthy baselines. Still, results from our small sample suggest there may be a role for the assessment of HRV in Lassa fever prognostication that can be defined with further investigation.

The shaded grey areas in Figure 3 represent periods of patient decompensation toward death that were captured by objective changes in vital signs, such as an acute increase in HR or RR. If used as real-time telemetry, the study’s monitoring system has the potential to identify these changes more rapidly than the current system of collecting patient vital signs every 30 minutes at the KGH Lassa ward, creating opportunities for earlier intervention and better outcomes^2^.

Although the use of wearable biosensor devices to remotely monitor individuals with severe acute Lassa fever appears to be feasible for both adults and children in Sierra Leone, further adoption poses several challenges. First, we encountered recurrent lapses in patient data collection due to failure of the devices’ adhesive backing, which was indicated by elevations in impedance detected by the sensor. Temperatures during the dry season in Sierra Leone frequently surpass 38 °C with a high relative humidity^24^. These climatic conditions, along with poor ventilation in patient rooms in the Lassa ward, likely led to patient perspiration causing occasional adhesive failure on the devices.

Second, due to the floor plan of the Lassa ward at KGH, Android devices were occasionally out of range for the monitoring devices’ Bluetooth connectivity, leading to further lapses in patient data collection. This was addressed by situating the Android devices as close as possible to the patient area, though they could ultimately not be brought into the patient area due to the location of power outlets and concerns about infection control. However, even when both devices were within the Bluetooth radius of 10 meters, there were still occasional connectivity problems, an issue that was present in a previous study conducted in a Rwandan emergency department ^2^. Third, we observed several spurious vital sign readings that would be physiologically impossible. These readings prompted us to discard patient data that was generated concomitantly, and raises concerns about rare but easily identifiable issues with device calibration. Fourth, at nearly US$170.00 each in 2024, the cost of wearable biosensor devices–not including the necessary Android devices to accompany them–may be prohibitive for many settings in which Lassa fever is endemic.

Lastly, Sierra Leone suffers from a significant shortage of clinicians, with only one physician per ten thousand population^25^. The country’s official Lassa ward does not have access to organ support equipment, such as mechanical ventilators or hemodialysis machines, and routine laboratory studies are performed infrequently. Even with the ability to identify patients at risk for death based on vital sign trends or acute decompensation, the burden on local physicians and the extreme scarcity of sufficient resources for meaningful intervention limits this technology from being implemented to its full potential.

## LIMITATIONS

Our study has several limitations. A more robust statistical analysis of our patients was limited by a small sample size, owing to the relative scarcity of diagnosed Lassa fever in Sierra Leone, which averaged just 8.4 cases annually from 2019 to 2023 (unpublished; from KGH records). Additionally, the majority (81.3%) of continuous physiological data we collected was ultimately discarded due to poor quality. There was also a bias toward younger individuals in our study, with the median age falling in the pediatric range. This is in accordance with existing epidemiologic data from Sierra Leone that suggest the highest incidence of antigenemic Lassa fever is observed in children and young adults^10^.

## CONCLUSIONS

Here we demonstrate that continuous vital signs monitoring of patients with Lassa fever can be achieved in resource-limited settings and may allow for the detection of early signs of clinical deterioration. Similar to what has been observed in bacterial sepsis, we found that HR, RR, and HRV may be related to outcome in acute Lassa fever. Further research with a larger sample size is needed to determine whether trends of these physiologic parameters can be harnessed for Lassa fever prognostication. To improve the effectiveness of data collection using wearable devices in similar settings to Sierra Leone, efforts should be focused on the improvement of the adhesive in the context of patient perspiration and to ensure that the biosensor and Android devices are able to remain within Bluetooth range without breaches in infection control. If this can be achieved, then wearable biosensors may have a role in real-time remote patient monitoring of critically ill patients that pose an infectious risk in low-resource settings.

## Data Availability

All data produced in the present study are available upon reasonable request to the authors

## ACKNOWLEDGEMENTS

The authors are grateful for the assistance of Laura Hughes and Shirlee Wohl for their guidance during the analysis of these data.

## REFERENCES

1. Baelani, I. et al. Availability of critical care resources to treat patients with severe sepsis or septic shock in Africa: a self-reported, continent-wide survey of anaesthesia providers. Crit. Care 15, R10 (2011).

2. Garbern, S. C. et al. Validation of a wearable biosensor device for vital sign monitoring in septic emergency department patients in Rwanda. Digit Health 5, 2055207619879349 (2019).

3. Steinhubl, S. R., Marriott, M. P. & Wegerich, S. W. Remote sensing of vital signs: A wearable, wireless band-aid’’ sensor with personalized analytics for improved Ebola patient care and worker safety. Glob Health Sci Pract 15, 516–519 (2015).

4. Shah, P. S. Wireless monitoring in the ICU on the horizon. Nature medicine vol. 26 316–317 (2020).

5. Edgcombe, H., Paton, C. & English, M. Enhancing emergency care in low-income countries using mobile technology-based training tools. Arch. Dis. Child. 101, 1149–1152 (2016).

6. Ahmad, S. et al. Continuous multi-parameter heart rate variability analysis heralds onset of sepsis in adults. PLoS One 4, e6642 (2009).

7. Steinhubl, S. R. et al. Validation of a portable, deployable system for continuous vital sign monitoring using a multiparametric wearable sensor and personalised analytics in an Ebola treatment centre. BMJ Glob Health 1, e000070 (2016).

8. Ngonzi, J., Boatin, A., Mugyenyi, G., Wylie, B. J. & Haberer, J. E. A functionality and acceptability study of wireless maternal vital sign monitor in a tertiary university teaching hospital in rural Uganda. J Womens Health Gyn 4, 1–8 (2017).

9. Ghiasi, S. et al. Sepsis Mortality Prediction Using Wearable Monitoring in Low-Middle Income Countries. Sensors 22, (2022).

10. Shaffer, J. G., et al. Lassa fever in post-conflict sierra leone. PLoS Negl. Trop. Dis. 8, e2748 (2014).

11. Murphy, H. L. & Ly, H. Pathogenicity and virulence mechanisms of Lassa virus and its animal modeling, diagnostic, prophylactic, and therapeutic developments. Virulence 12, 2989–3014 (2021).

12. UNDP (United Nations Development Programme). Human Development Report 2021-22: Uncertain Times, Unsettled Lives: Shaping Our Future in a Transforming World. (New York, 2022).

13. Henning, A. & Krawiec, C. Sinus Tachycardia. in StatPearls (StatPearls Publishing, Treasure Island (FL), 2023).

14. van den Berg, M. P., Hassink, R. J., Baljé-Volkers, C. & Crijns, H. Role of the autonomic nervous system in vagal atrial fibrillation. Heart 89, 333–335 (2003).

15. Parker, M. M., Shelhamer, J. H., Natanson, C., Alling, D. W. & Parrillo, J. E. Serial cardiovascular variables in survivors and nonsurvivors of human septic shock: heart rate as an early predictor of prognosis. Crit. Care Med. 15, 923–929 (1987).

16. Fleming, S. et al. Normal ranges of heart rate and respiratory rate in children from birth to 18 years of age: a systematic review of observational studies. Lancet 377, 1011–1018 (2011).

17. Garry, R. F. Lassa Fever: Epidemiology, Immunology, Diagnostics, and Therapeutics. vol. 440 165–192 (Springer International Publishing, 2023).

18. Duvignaud, A. et al. Lassa fever outcomes and prognostic factors in Nigeria (LASCOPE): a prospective cohort study. The Lancet Global Health 9, e469–e478.

19. Chen, W.-L. et al. Heart rate variability measures as predictors of in-hospital mortality in ED patients with sepsis. Am. J. Emerg. Med. 26, 395–401 (2008).

20. Pontet, J. et al. Heart rate variability as early marker of multiple organ dysfunction syndrome in septic patients. J. Crit. Care 18, 156–163 (2003).

21. Quinten, V. M., van Meurs, M., Renes, M. H., Ligtenberg, J. J. M. & Ter Maaten, J. C. Protocol of the sepsivit study: a prospective observational study to determine whether continuous heart rate variability measurement during the first 48 hours of hospitalisation provides an early warning for deterioration in patients presenting with infection or sepsis to the emergency department of a Dutch academic teaching hospital. BMJ Open 7, e018259 (2017).

22 Task Force of the European Society of Cardiology the North American Society of Pacing Electrophysiology. Heart rate variability: standards of measurement, physiological interpretation, and clinical use Circulation (1996).

23. Piepoli, M., Garrard, C. S., Kontoyannis, D. A. & Bernardi, L. Autonomic control of the heart and peripheral vessels in human septic shock. Intensive Care Med. 21, 112–119 (1995).

24. Simbo, R. T., Gogra, A. B., Kawa, Y. K. & Moiwo, P. J. Seasonal Effect on Weather Elements on Water Table Fluctuation in Potable Wells in Kono District, Eastern Sierra Leone. Open Journal of Applied Sciences 13, 2198– 2209 (2023).

25. World Health Organization. The National health Workforce Accounts database. https://apps.who.int/nhwaportal, https://www.who.int/activities/improving-health-workforce-data-and-evidence.

